# Combining VEGFR tyrosine kinase inhibitors and PD-1/PD-L1 inhibitors versus VEGFR tyrosine kinase inhibitors monotherapy in renal cell carcinoma: a target trial emulation

**DOI:** 10.64898/2026.06.30.26356941

**Authors:** Daling Shi, Xiaochun Li, Yao Chen, Yijiang Chen, Qianqian Song, Jing Su

## Abstract

**Importance:** Combinations of VEGFR tyrosine kinase inhibitors (TKIs) and immune checkpoint inhibitors (ICIs), such as antibodies to programmed cell death-1 (PD-1), or to its ligand PD-L1, are now first-line standard of care for renal cell carcinoma (RCC), but the pivotal clinical trials excluded patients with common comorbidities, leaving their real-world effectiveness uncertain.

**Objective:** To determine whether adding PD-1/PD-L1 inhibitors to VEGFR-TKIs therapy is associated with improved overall survival in a real-world RCC cohort.

**Design, Setting, and Participants:** This retrospective cohort study used a target trial emulation framework and real-world electronic health records data from the University of Florida Health Integrated Data Repository (IDR). Data was analyzed from September 2009 through June 2023. Adult patients (≥18 years) with confirmed RCC and at least one VEGFR-TKIs prescription were eligible. The date of the first VEGFR-TKIs prescription was defined as the index date, and patients were followed for up to 24 months. Variable-ratio propensity score matching (up to 2:1) across 13 baseline covariates was used to emulate randomized treatment assignments. Of 107,783 patients screened, 387 met eligibility criteria, and 319 remained in the matched cohort.

**Exposures:** VEGFR-TKIs monotherapy (control group) versus VEGFR-TKIs combined with PD-1/PD-L1 inhibitors (experimental group).

**Main Outcomes and Measures:** Overall survival (OS), analyzed by weighted Kaplan-Meier estimation, cluster-robust Cox regression, and restricted mean survival time (RMST) at τ = 24 months, prespecified given anticipated non-proportional hazards.

**Results:** Among 319 matched patients (mean [SD] age, 62 [12] years; 76% male), 107 deaths occurred (33.5%). Twelve-month OS was higher in the combination arm (81.8%; 95% CI, 74.7–89.6%) than VEGFR-TKIs monotherapy (68.1%; 95% CI, 61.1–76.0%), converging by 24 months (61.1% vs 56.7%). The Cox hazard ratio was 0.718 (95% CI, 0.484– 1.064; *P* = 0.0986). RMST was 2.79 months greater with combination therapy (95% CI, 0.93–4.65; *P* = 0.0033).

**Conclusions:** Adding PD-1/PD-L1 inhibitors to VEGFR-TKIs therapy was associated with a statistically significant and clinically meaningful gain in restricted mean survival, supporting the real-world generalizability of combination therapy and the importance of appropriate treatment effect measures under non-proportional hazards.

## INTRODUCTION

Renal cell carcinoma (RCC) accounts for approximately 81,800 new diagnoses and more than 14,000 deaths in the United States each year, with a substantial proportion of patients presenting with locally advanced or metastatic disease at the time of diagnosis.^1^ Over the past decade, treatment for RCC has changed greatly because of drugs that target the vascular endothelial growth factor receptor (VEGFR) pathway and the programmed cell death protein 1 / programmed death-ligand 1 (PD-1/PD-L1) immune checkpoint axis.^2,3^ Multiple pivotal randomized clinical trials have shown that combining a VEGFR tyrosine kinase inhibitor (TKI) with a PD-1/PD-L1 inhibitor improves outcomes in RCC. These trials include pembrolizumab plus axitinib (KEYNOTE-426)^4^, avelumab plus axitinib (JAVELIN Renal 101)^5^, nivolumab plus cabozantinib (CheckMate 9ER)^6^, and lenvatinib plus pembrolizumab (CLEAR)^7^. All of these regimens showed better overall or progression-free survival than VEGFR-TKIs monotherapy, and they are now recommended as first-line options in clinical guidelines. However, these trials were conducted in carefully selected patient populations and under controlled study conditions. Eligibility criteria often require good performance status, and trial protocols apply close monitoring and standardized treatment that may not reflect routine care. As a result, patients seen in everyday practice tend to be older and more heterogeneous, with a wider range of comorbidities, disease severity, and prior treatments. Because of this, it is not clear whether the benefits observed in trials can be fully generalized to everyday clinical practice. This question matters because treatment decisions are ultimately made in real clinical settings, and understanding how well these therapies work in practice is important for patients, clinicians, and healthcare systems.^8^

Real-world data can help address this gap. In routine practice, patients differ widely in demographic characteristics, comorbidities, disease severity, prior treatment, and overall clinical condition. In addition, physicians do not assign treatments randomly. Instead, treatment choice is shaped by patient characteristics and clinical judgment. For these reasons, the outcomes observed in real-world practice may differ from those reported in clinical trials.^9^ Evaluating treatment effectiveness in real-world clinical settings provides evidence that complements traditional clinical trial results and better represents the patients treated in routine practice.^10^ The need for real-world effectiveness evidence is particularly important for first-line combination therapies in advanced RCC, as the pivotal clinical trials supporting these regimens have only recently reached maturity. KEYNOTE-426 and JAVELIN Renal 101 have recently completed follow-up, while CheckMate 9ER and CLEAR remain ongoing. Although these trials established the efficacy of combination therapy under controlled study conditions, their findings have not yet been extensively validated in broader real-world populations. At the same time, increasing numbers of patients have now received these regimens in routine practice, creating sufficiently large real-world cohorts for analysis. This provides a timely opportunity to leverage observational data through target trial emulation (TTE), reproducing key features of the landmark randomized clinical trials while generating effectiveness estimates in more representative patient populations and assessing the real-world generalizability of trial findings.

Although real-world data are valuable, traditional observational studies are susceptible to selection bias and confounding, which can weaken causal conclusions.^11^ Target trial emulation (TTE) is an analytic framework designed to strengthen causal inference from real-world observational data by explicitly emulating the protocol of a hypothetical randomized controlled trial.^12,13^ By prespecifying eligibility criteria, treatment strategies, outcomes, follow-up rules, and statistical analyses in a way similar to the randomized clinical trials protocol, TTE helps reduce several common sources of bias in electronic health record (EHR)-based studies. These include immortal time bias, prevalent user bias, and confounding by indication, which have historically affected the validity of real-world analyses.^13,14^ Propensity score methods, applied within the TTE framework, further approximate the balanced covariate distributions achieved by randomization and allow for more credible estimation of causal treatment effects from observational data.^15^ TTE-based analyses have been shown to produce effect estimates that are directionally consistent with randomized clinical trial results across multiple therapeutic areas, supporting their use as a rigorous complement to experimental evidence when trial data are unavailable or limited in generalizability to real-world populations.^16^

Most previous real-world emulations of the pivotal RCC clinical trials^17-20^ have typically emulated a single trial, so they were limited to one specific two-or three-drug regimen and were less able to represent the wider class of agents used in practice. In this study, we take a broader view: rather than focusing on a single named combination, we encompass the commonly used VEGFR-TKIs and PD-1/PD-L1 inhibitors as therapeutic classes, which better reflects the variety of agents prescribed in routine care. We applied a TTE framework to real-world data from the University of Florida Health Integrated Data Repository (UF Health IDR)^21^, a secure clinical data warehouse that consolidates EHR and administrative data across UF Health and structures them using the PCORnet Common Data Model. Within this framework, we used variable-ratio propensity score matching to emulate the balanced treatment assignment of a randomized trial and compared VEGFR-TKIs monotherapy with VEGFR-TKIs plus PD-1/PD-L1 inhibitors combination therapy. The aim of this study was to determine whether the addition of PD-1/PD-L1 inhibitors to VEGFR-TKIs therapy is associated with an overall survival benefit in a real-world RCC cohort.

## RESULTS

### Patient characteristics

Among the 107,783 patients in UF Health with prescribing records and medical encounters between September 21, 2009, and June 22, 2023, 1,436 were identified as eligible patients with VEGFR-TKIs exposure after excluding 106,347 individuals without VEGFR-TKIs exposure (only PD-1/PD-L1 inhibitors: n = 2,127; neither drug: n = 104,220). After excluding 1,046 patients without RCC, 390 RCC patients with VEGFR-TKIs exposure were identified. Of the 390 RCC patients, 267 initiated VEGFR-TKIs monotherapy and 123 initiated VEGFR-TKIs combined with PD-1/PD-L1 inhibitors, with the first VEGFR-TKIs treatment date defined as the index date. After excluding 3 patients with invalid survival records due to data inconsistencies (e.g., death dates preceding medication order dates), the study cohort consisted of 387 patients (VEGFR-TKIs monotherapy arm [control/CTRL group]: n = 264, VEGFR-TKIs with PD-1/PD-L1 inhibitors arm [experimental/EXP group]: n = 123). Of these, 130 patients (33.6%) experienced the primary endpoint of all-cause mortality, while 257 (66.4%) were censored. After variable ratio propensity score matching (PSM), 204 individuals were included in the CTRL group and 115 in the EXP group for outcome analyses. Among matched pairs, 89 subclasses achieved 1:2 matching and 26 achieved 1:1 matching, yielding a control-to-treatment ratio of approximately 1.77:1. The study cohort construction and exclusion criteria are shown in **Figure 1**.

**Figure 1.**
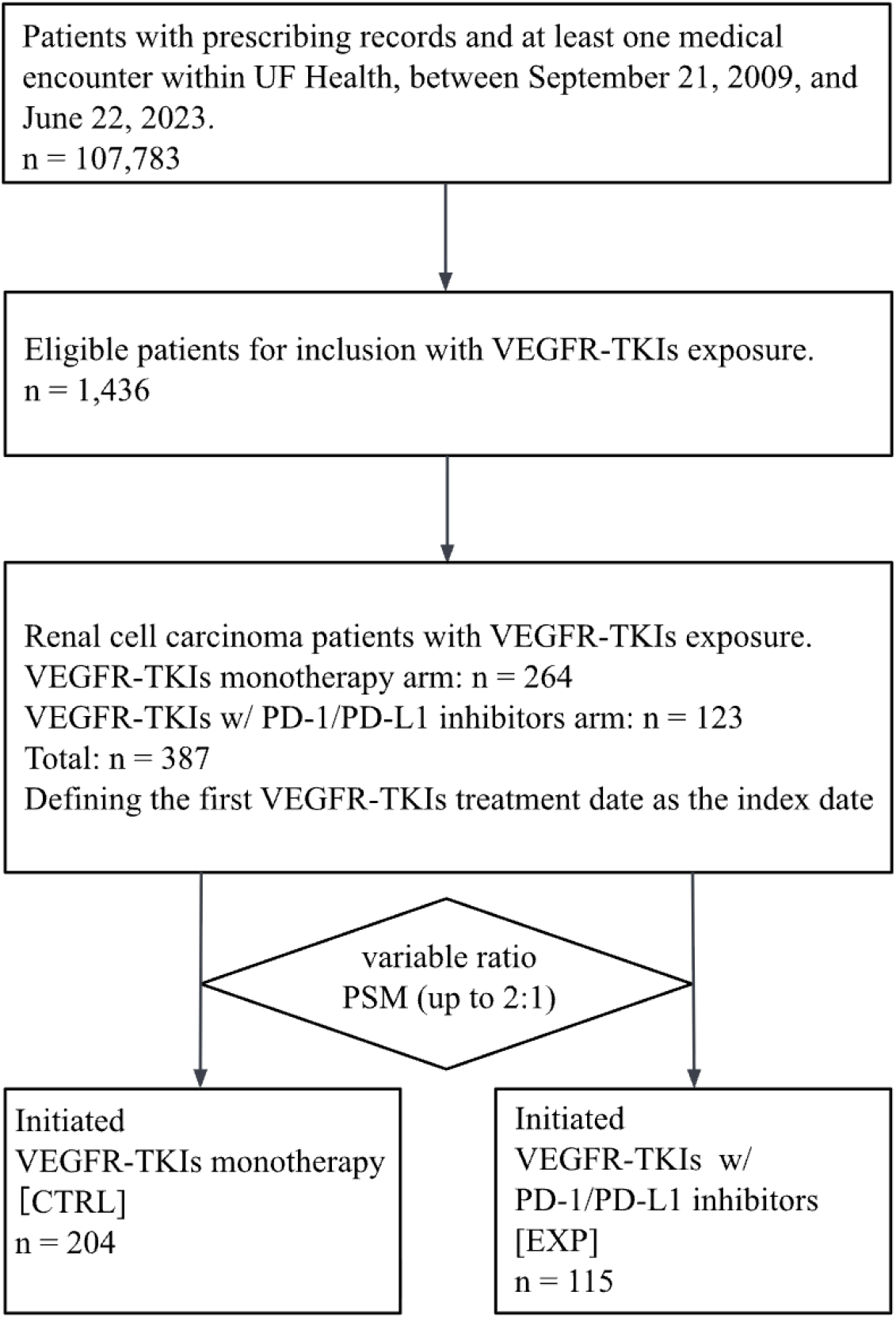
Study cohort construction and inclusion criteria. Flow diagram of cohort selection from the UF Health Integrated Data Repository. VEGFR-TKIs, vascular endothelial growth factor receptor tyrosine kinase inhibitors; PD-1, programmed cell death protein 1; PD-L1, programmed death-ligand 1; PSM, propensity score matching; CTRL, control; EXP, experimental.

In the study cohort, the EXP group was younger on average (mean [SD] age, 61.9 [12.0] years) than the CTRL group (63.3 [11.9] years) and included a higher proportion of male patients (EXP: 80.5% vs. CTRL: 72.0%). Most patients in both groups were White (EXP: 87.0%; CTRL: 79.9%), and Non-Hispanic Black patients constituted 8.9% and 10.6% of the EXP and CTRL groups, respectively. Within the 24-month administrative censoring window, 39 deaths (31.7%) were recorded in the EXP group and 91 deaths (34.5%) in the CTRL group. To minimize confounding between the VEGFR-TKIs alone and VEGFR-TKIs + PD-1/PD-L1 inhibitors groups, variable ratio propensity score matching (PSM) was performed across 13 baseline covariates, including demographic characteristics and key comorbidities. Prior to matching, the most notable imbalances were observed in chronic kidney disease (CKD; SMD = 0.316), liver disease (LD; SMD = 0.227), myocardial infarction (MI; SMD = 0.226), cerebrovascular disease (CVD; SMD = 0.225), and sex distribution (SMD = 0.201). After PSM, covariate balance was substantially improved across all 13 variables, with post-matching SMDs ranging from 0.0004 to 0.0924, all below the pre-specified threshold of 0.10 (**Table 1**). **Figure 2** illustrates and confirms this improvement.

**Table 1.** Baseline characteristics before and after propensity score matching. Demographic, behavioral, and comorbidity characteristics of patients initiating VEGFR-TKIs monotherapy (CTRL) versus VEGFR-TKIs + PD-1/PD-L1 inhibitors (EXP), shown for the full and matched cohorts. Categorical variables are reported as counts and percentages, n (%), and age is reported as mean (SD). Cells with counts below 20 are masked (<20) in accordance with institutional small-cell privacy policy. between-group balance is assessed by the standardized mean difference (SMD), with SMD < 0.10 indicating adequate balance. After matching, all 13 covariates fell below this threshold. Abbreviations: SMD, standardized mean difference; SD, standard deviation; CTRL, control; EXP, experimental.

| Variable | Before Matching |  |  | After Matching |  |  |
| --- | --- | --- | --- | --- | --- | --- |
|  | CTRL, n (%) | EXP, n (%) | SMD | CTRL, n (%) | EXP, n (%) | SMD |
| n | 264 | 123 | - | 204 | 115 | - |
| Sex (Male) | 190 (72.0) | 99 (80.5) | 0.201 | 152 (74.5) | 92 (80.0) | 0.0924 |
| Smoking | 153 (58.0) | 76 (61.8) | 0.078 | 122 (59.8) | 70 (60.9) | 0.0266 |
| Myocardial Infarction (MI) | 25 (9.5) | 21 (17.1) | 0.226 | 23 (11.3) | <20 (<17.4) | 0.0258 |
| Congestive Heart Failure (CHF) | 53 (20.1) | 30 (24.4) | 0.104 | 44 (21.6) | 26 (22.6) | 0.0209 |
| Peripheral Vascular Disease (PVD) | 38 (14.4) | <20 (<16.3) | 0.170 | <20 (<9.8) | <20 (<17.4) | 0.0136 |
| Cardiovascular Disease (CVD) | 50 (18.9) | 35 (28.5) | 0.225 | 45 (22.1) | 28 (24.3) | 0.0206 |
| Chronic Obstructive Pulmonary Disease (COPD) | 40 (15.2) | 20 (16.3) | 0.031 | 31 (15.2) | <20 (<17.4) | 0.0239 |
| Paralysis (PARA) | 21 (8.0) | <20 (<16.3) | 0.007 | <20 (9.8) | <20 (<17.4) | 0.0320 |
| Diabetes | 95 (36.0) | 51 (41.5) | 0.113 | 80 (39.2) | 47 (40.9) | 0.0089 |
| Chronic Kidney Disease (CKD) | 122 (46.2) | 76 (61.8) | 0.316 | 111 (54.4) | 68 (59.1) | 0.0177 |
| Liver Disease (LD) | 89 (33.7) | 55 (44.7) | 0.227 | 80 (39.2) | 47 (40.9) | 0.0090 |
| Non-Hispanic Black (ETH_BLACK) | 28 (10.6) | <20 (<16.3) | 0.056 | 20 (9.8) | <20 (<17.4) | 0.0146 |
| Age (mean (SD)) | 63.28 (11.85) | 61.91 (11.97) | 0.115 | 62.27 (11.85) | 62.03 (12.10) | 0.0004 |

**Figure 2.**
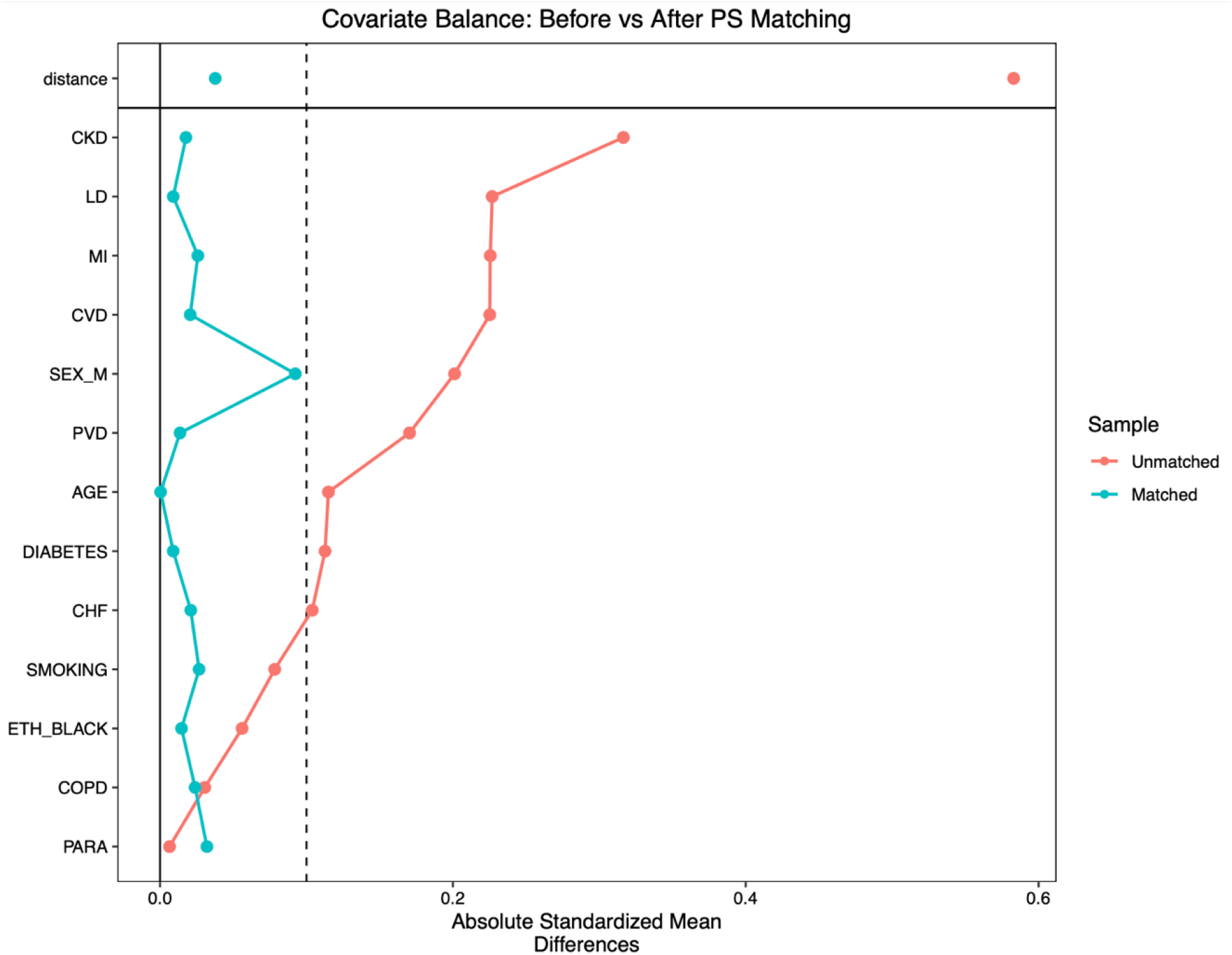
Covariate balance before and after propensity score matching. Love plot showing absolute SMD for each baseline covariate in the unmatched (red) and matched (blue) cohorts. Each point represents one covariate; the propensity score (distance) is shown separately above the horizontal line. The dashed vertical line at 0.1 denotes the conventional threshold for acceptable balance. Before matching, several covariates exceeded the 0.1 threshold, and the propensity score showed marked imbalance. After variable-ratio propensity score matching, the SMD for the propensity score and for nearly all covariates fell below 0.1, indicating substantially improved balance between the VEGFR-TKIs monotherapy and combination therapy arms. SMD, standardized mean difference.

### Compared VEGFR-TKIs monotherapy and VEGFR-TKIs with PD-1/PD-L1 inhibitors on the outcome of interests

To assess the comparative survival benefit of VEGFR-TKIs with PD-1/PD-L1 inhibitors (EXP) group versus VEGFR-TKIs monotherapy (CTRL) group, Kaplan-Meier (KM) analysis of overall survival (OS) was conducted in the study cohort after propensity score matching, which comprised 204 patients in the CTRL group and 115 patients in the EXP group (total n = 319). Over the entire follow-up period, 107 deaths were recorded, yielding an overall event rate of 33.5%. The relatively low event rate, combined with the fact that median OS was not reached in either group, indicates that a substantial proportion of patients in both cohorts remained alive at the time of data cutoff. Despite the median OS remaining undefined for both groups, the lower bound of the 95% confidence interval (CI) for median OS in the CTRL group was estimated at 20.6 months, providing a conservative anchor for the survival trajectory of this cohort. In the EXP group, the 95% confidence interval for median OS could not be estimated because there were few late events and many patients had censored survival times. Although this limited the estimation of median OS, it reflects the generally good prognosis of the study cohort and/or the relatively short follow-up period. Kaplan-Meier survival curves with risk tables for the study cohort are presented in **Figure 3**.

**Figure 3.**
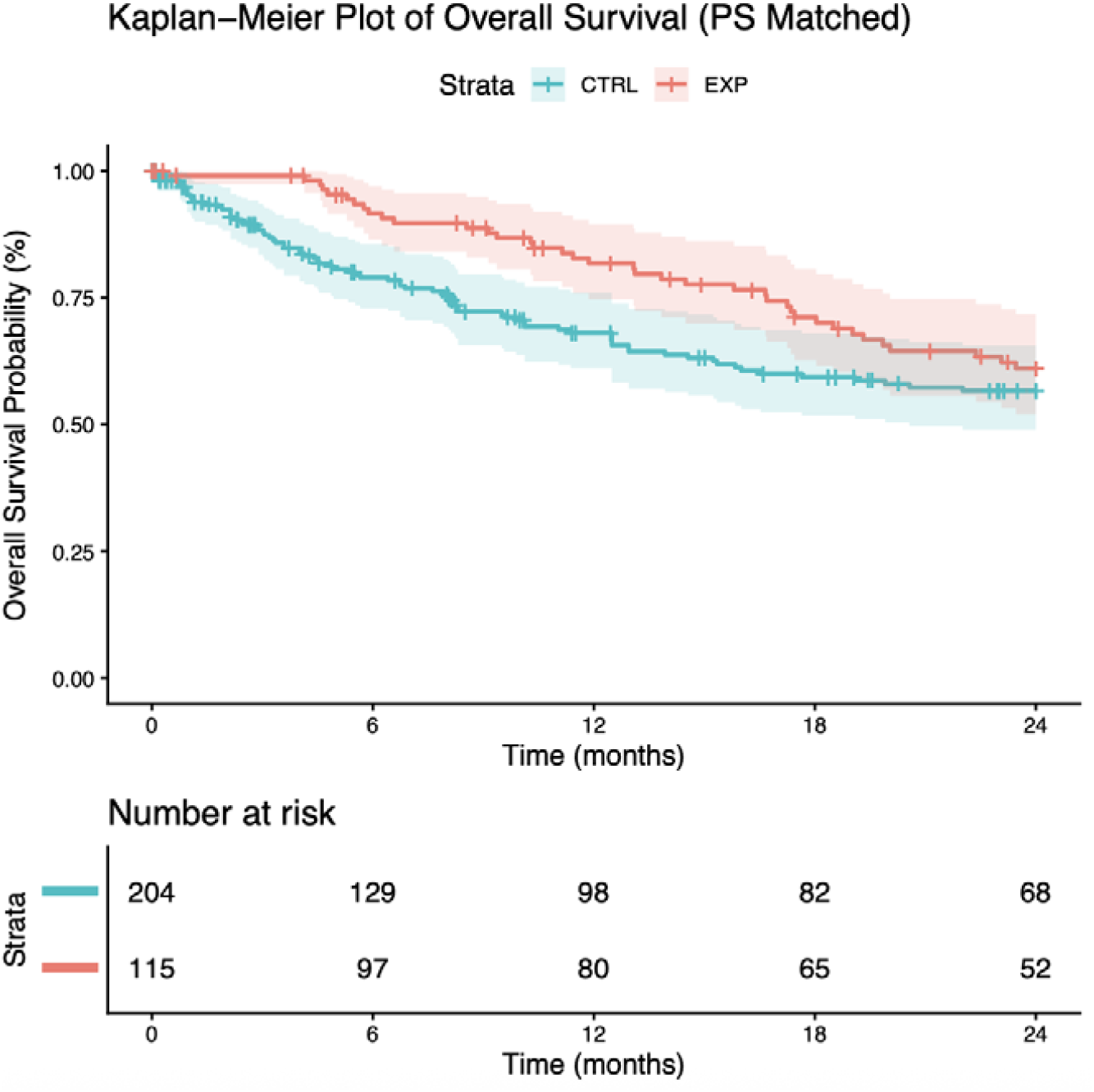
Overall survival in the study cohort. Kaplan–Meier curves for overall survival in the study cohort, comparing patients initiating VEGFR-TKIs monotherapy (CTRL, blue) with those initiating VEGFR-TKIs plus PD-1/PD-L1 inhibitors combination therapy (EXP, red). Shaded bands represent 95% confidence intervals and tick marks denote censored observations. The combination therapy arm showed higher survival probability than the monotherapy arm over 24 months of follow-up (*P* = 0.0986, weighted Cox proportional hazards model). The number of patients at risk in each arm is shown below the plot. CTRL, control (monotherapy); EXP, experimental (combination therapy); PS, propensity score; VEGFR-TKIs, vascular endothelial growth factor receptor tyrosine kinase inhibitors; PD-1, programmed cell death protein 1; PD-L1, programmed death-ligand 1; ICIs, immune checkpoint inhibitors.

To quantify survival differences at clinically meaningful time points, analyses were performed at 12 and 24 months (**Table 2**). At 12 months, the EXP group showed a markedly higher OS rate of 81.8% (95% CI, 74.7–89.6%) compared with 68.1% (95% CI, 61.1–76.0%) in the CTRL group, representing an absolute difference of 13.7 percentage points. This early separation of the survival curves provides preliminary evidence of a statistically meaningful early survival benefit associated with the combination of VEGFR-TKIs and PD-1/PD-L1 inhibitors. At 24 months, OS rates were 61.1% (95% CI, 52.0–71.7%) and 56.7% (95% CI, 49.0–65.6%) in the EXP and CTRL groups, respectively. Although the absolute difference narrowed to 4.4 percentage points and the confidence intervals began to overlap, the EXP group still showed a numerically higher survival probability, suggesting a durable but attenuated benefit over the medium term. The pattern of early separation followed by gradual convergence of the KM curves is consistent with a treatment effect that is strongest in the early post-treatment period.^22^ This may reflect the rapid antiangiogenic and tumor-stabilizing effects of VEGFR-TKIs, together with the more delayed and heterogeneous benefit of PD-1/PD-L1 blockade, as well as the eventual emergence of resistance and disease progression over time.^23^

**Table 2.** Kaplan-Meier Survival Estimates and Cox Proportional Hazards Analysis of Overall Survival in the Propensity Score Matched Cohort. Survival outcomes were estimated in a propensity score matched cohort of 319 patients. Events represent inverse probability-weighted death counts. Median Overall Survival (OS) was not reached (NR) in either group during the observation period. Overall survival (OS) rates at 12 and 24 months are presented with 95% confidence intervals (CIs) derived from Greenwood’s formula. The hazard ratio (HR) and corresponding 95% CI were estimated using Cox proportional hazards regression. Abbreviations: CI, confidence interval; HR, hazard ratio; mo, months; NR, not reached; OS, overall survival.

|  | VEGFR-TKIs monotherapy (CTRL) group | VEGFR-TKIs with PD-1/PD-L1 inhibitors (EXP) group |
| --- | --- | --- |
| **n** | 204 | 115 |
| **Events** | 70.1 | 38.0 |
| **Median OS (95% CI)** | NR (20.6–NR) | NR (NR–NR) |
| **12-mo OS rates (95% CI)** | 68.1% (61.1–76.0%) | 81.8% (74.7–89.6%) |
| **24-mo OS rates (95% CI)** | 56.7% (49.0–65.6%) | 61.1% (52.0–71.7%) |
| **Cox HR (95% CI)** | — | 0.718 (0.484–1.064) |
| **Cox P-value** | — | 0.0986 |

A Cox proportional hazards regression model with robust variance estimation clustered by matched subclass was performed to compare overall survival between groups while accounting for the matched study design **(Table 2)**. This model yielded a hazard ratio (HR) of 0.718 (95% CI, 0.484–1.064; *P* = 0.0986), corresponding to a 28.2% relative reduction in the instantaneous hazard of death for patients receiving VEGFR-TKIs with PD-1/PD-L1 inhibitors compared with VEGFR-TKIs monotherapy. The point estimate, effect direction, and overall pattern of the survival curves were consistent across all analytical approaches, including KM estimation (**Figure 3**) and Cox regression (**Table 2**), supporting the internal consistency of the observed findings. However, formal evaluation of the proportional hazards (PH) assumption using scaled Schoenfeld residuals revealed significant time-dependency of the treatment effect (χ^2^ = 15.2, df = 1, *P* = 9.7 × 10^−5^), indicating that the hazard ratio was not constant over the follow-up period and that the Cox model may therefore underestimate the magnitude of early treatment benefit. So we estimated treatment effects using restricted mean survival time (RMST) at a pre-specified truncation time of τ = 24 months, an approach that is robust to non-proportional hazards and provides a clinically interpretable measure of absolute survival benefit without assuming a constant hazard ratio (**Table 3**). The mean survival time within 24 months was 19.70 months (95% CI, 18.42–20.97) in the EXP group compared with 16.91 months (95% CI, 15.56–18.27) in the CTRL group. The between-group difference in RMST was 2.79 months (95% CI, 0.93–4.65; *P* = 0.0033), representing a statistically significant and clinically meaningful absolute gain of EXP group in survival time, compared with the CTRL group. The RMST ratio was 1.17 (95% CI, 1.05–1.29; *P* = 0.0037), and the restricted mean time lost (RMTL) ratio was 0.61 (95% CI, 0.43–0.86; *P* = 0.0055), collectively corroborating the survival advantage observed in patients receiving combination therapy.

**Table 3.** Proportional hazards assumption test and restricted mean survival time (RMST) analysis in the propensity score-matched cohort. Section A presents the results of the proportional hazards assumption test performed using scaled Schoenfeld residuals, reporting the chi-squared statistic (χ^2^), degrees of freedom (df), and associated *P*-value for the treatment variable. Section B reports arm-specific restricted mean survival time (RMST) estimates with 95% confidence intervals at a pre-specified truncation time of τ = 24 months; between-group *P*-values are not applicable for individual arm estimates and are therefore denoted by dashes. Section C presents the between-group contrast, including the absolute difference in RMST and the RMST ratio, each with 95% confidence intervals and two-sided *P*-values. EXP, experimental group (VEGFR-TKIs combined with PD-1/PD-L1 inhibitors); CTRL, control group (VEGFR-TKIs alone).

|  | EXP | CTRL | P-value |
| --- | --- | --- | --- |
| **A. Proportional hazards assumption test (Schoenfeld residuals)** |  |  |  |
| Treatment (EXP vs. CTRL) | $\chi^2 = 15.2$ | $df = 1$ | $9.7 \times 10^{-5}$ |
| **B. Restricted mean survival time by arm ($\tau = 24$ months)** |  |  |  |
| RMST, months (95% CI) | 19.70 (18.42–20.97) | 16.91 (15.56–18.27) | — |
| **C. Between-group contrast** |  |  |  |
|  | **Estimate** | **95% CI** | **P-value** |
| RMST difference (EXP – CTRL), months | 2.79 | 0.93–4.65 | 0.0033 |
| RMST ratio (EXP / CTRL) | 1.17 | 1.05–1.29 | 0.0037 |

## DISCUSSION

In this real-world cohort study of RCC patients, the combination of VEGFR-TKIs therapy with PD-1/PD-L1 inhibitors was associated with a statistically significant improvement in overall survival compared with VEGFR-TKIs therapy alone, as demonstrated by the RMST analysis. The observed 2.79-month gain in restricted mean survival over 24 months represents a clinically meaningful absolute benefit in a population with a progressive and frequently fatal disease course. Although the conventional Cox hazard ratio was 0.718 (95% CI, 0.484–1.064; P = 0.0986), testing indicated evidence of non-proportional hazards (χ^2^ = 15.2; P = 9.7 × 10^−5^), suggesting that the hazard ratio alone may not adequately summarize the treatment effect over time. The RMST, which does not rely on the proportional hazards assumption, provides a more valid and clinically interpretable estimate of the treatment difference in this context. The time-varying treatment effect in our data shows an early survival benefit with the combination therapy, with higher 12-month OS (81.8% vs. 68.1%), which then narrows by 24 months (61.1% vs. 56.7%). This pattern is consistent with the complementary mechanisms of VEGFR tyrosine kinase inhibition and PD-1/PD-L1 blockade. The early separation of the survival curves may be driven by the rapid anti-angiogenic and tumor-stabilizing effects of VEGFR-TKIs. In contrast, PD-1/PD-L1 blockade works more gradually by activating T-cell-mediated anti-tumor immunity, which may help maintain benefit over the medium term. The subsequent convergence likely reflects the eventual emergence of resistance and disease progression. This pattern, with a strong early benefit that attenuates but does not disappear, has been widely documented in randomized clinical trials of ICI-based combinations for advanced RCC^3,24^. Specifically, in the JAVELIN Renal 101 trial, early crossing of the survival curves was observed before avelumab plus axitinib conferred sustained OS benefit over sunitinib (HR, 0.78; 95% CI, 0.554–1.084)^19^. Similarly, CheckMate 214 demonstrated that the OS benefit of nivolumab plus ipilimumab became more pronounced with longer follow-up, a pattern attributed to the durable disease control characteristic of ICI-based regimens^25^. Our real-world RMST estimate of a 2.79-month gain over 24 months, corresponding to an approximately 16% increase in time alive, is directionally and quantitatively consistent with these established trial results.

To our knowledge, this study is among the first real-world studies from a United States academic medical center to directly compare VEGFR-TKIs monotherapy versus VEGFR-TKIs + PD-1/PD-L1 inhibitors combination therapy in RCC patients using time-to-event methodology with propensity score adjustment. The RMST ratio of 1.165 implies that patients receiving combination therapy spent 16.5% more of the 24-month window alive, a gain that is likely to be perceived as beneficial by patients and clinicians. The RMTL ratio of 0.61 further underscores that the combination arm experienced substantially less time lost to death during follow-up. The elevated comorbidity burden in the EXP group at baseline, including higher rates of CKD (61.8% vs. 46.2%), liver disease (44.7% vs. 33.7%), CVD (28.5% vs. 18.9%), and MI (17.1% vs. 9.5%) before matching, argues against a healthy-user bias explanation for the observed benefit. Had the combination therapy group been systematically healthier, we would expect these imbalances to favor EXP; instead, the EXP group carried a greater comorbidity burden. After PSM successfully balanced all covariates to an SMD < 0.10, the persistence of a survival advantage in the EXP arm strengthens the causal interpretation of the treatment effect. This finding is consistent with real-world evidence from other settings demonstrating that the survival benefit of VEGFR-TKIs + PD-1/PD-L1 inhibitors combinations is maintained in patients with comorbidities who may not have met strict RCT eligibility criteria.

This study has several methodological strengths. First, we employed variable ratio nearest-neighbor PSM (up to 2:1), which retained a substantially larger proportion of eligible controls than fixed 1:1 matching would allow. It is an important consideration in a cohort where controls outnumbered treated patients approximately 2:1 before matching. The imposed caliper of 0.2 standard deviations^26^ on the propensity score logit adheres to the established threshold for preventing poor-quality matches while maintaining adequate sample retention. Second, we used cluster-robust standard errors with clustering at the matched subclass level. This is important in variable-ratio matching because it accounts for within-subclass correlation. Without it, confidence intervals would be too narrow and error could be inflated. Third, RMST was pre-specified in the statistical analysis plan as the primary effect measure if the proportional hazards assumption was not met, avoiding data-driven selection of effect measures. In addition, RMST provides a clear advantage by expressing the treatment effect as an absolute gain in survival time. It is easy to interpret clinically, does not rely on the proportional hazards assumption required by the Cox model, and is directly meaningful for both patients and clinicians. Fourth, the comorbidity assessment used each patient’s full diagnostic history rather than a fixed lookback period. This reduces the chance of missing or misclassifying chronic conditions that are recorded intermittently, which is a known source of residual confounding in EHR-based propensity score models. Finally, mortality outcomes were obtained from the UF Health Tumor Registry rather than relying only on EHR records. Tumor registry data follow standardized collection and quality control procedures, which improves the completeness and reliability of death ascertainment. This reduces the likelihood of outcome misclassification and strengthens the validity of survival analyses compared with using routine clinical records alone.

Several limitations warrant consideration. As a retrospective observational study using real-world data rather than randomized treatment assignment, residual confounding cannot be completely excluded, even after propensity score matching and the overall good balance achieved after matching. In particular, several clinically important variables were unavailable or incompletely captured in the EHR, including ECOG performance status, IMDC risk category, tumor histology, line of therapy, and other factors that may influence both treatment selection and survival outcomes. Furthermore, treatment heterogeneity within the combination arm (varying VEGFR tyrosine kinase inhibitors and PD-1/PD-L1 inhibitors agents, sequences, dosage, and durations) limits regimen-specific interpretation. The modest sample size constrains the precision of subgroup analyses, and EHR-based mortality ascertainment may incompletely capture deaths occurring outside the UF Health system. In addition, the 24-month administrative censoring window may not reflect the full long-term survival benefit of immunotherapy, and the absence of biomarker data (PD-L1 expression, genomic profiling, treatment doses) precludes biomarker-stratified analyses. Finally, although comorbidity assessment followed the NCI Comorbidity Index framework to improve accuracy and consistency, ICD-based coding may not fully capture the complete burden of chronic conditions in real-world clinical practice, as some comorbidities may be inconsistently recorded or missed in routine care data.

In conclusion, this real-world effectiveness study provides statistically significant and clinically meaningful evidence that the addition of PD-1/PD-L1 inhibitors to VEGFR-TKIs therapy is associated with a 2.79-month gain in restricted mean overall survival over 24 months compared with VEGFR-TKIs therapy alone in RCC patients. The non-proportional hazards pattern observed is consistent with the early antiangiogenic benefit of VEGFR-TKIs followed by the gradual development of durable immunotherapeutic response, and RMST analysis provides a valid and informative effect summary in this setting. These findings support the real-world generalizability of combination therapy regimens, and underscore the importance of selecting appropriate statistical treatment effect measures when evaluating ICI-based therapies. Larger multi-center real-world datasets and longer follow-up will be necessary to further characterize the long-term benefit, identify predictive biomarkers, and optimize regimen selection in this patient population.

## METHODS

### Data source

This retrospective observational cohort study used real-world electronic health record (EHR) data from the University of Florida Health Integrated Data Repository (UF Health IDR). Ethical approval was granted by the University of Florida Institutional Review Board (IRB202601021), and the requirement for informed consent was waived given the retrospective study design and use of de-identified patient data. Data were obtained from the UF Health IDR, a secure clinical data warehouse that consolidates information from multiple UF Health clinical and administrative platforms^21^. The repository encompasses more than one billion clinical observations from over two million patients, providing the scale necessary for population-level research aimed at advancing scientific discovery and improving patient care. Data were structured according to the PCORnet Common Data Model (CDM), and six source tables were used in this analysis: PRESCRIBING, which captured medication orders and prescription dates; TUMOR_REGISTRY, which contained cancer diagnoses and age at diagnosis; DIAGNOSIS, which provided comorbidity codes drawn from the full longitudinal patient history; DEMOGRAPHIC, which included sex, race, ethnicity, and date of birth; DEATH, which recorded mortality outcomes; and ENCOUNTER, which documented healthcare visit dates. Drug identification was performed using RxNorm Concept Unique Identifiers (RxCUI) derived from standardized reference tables compiled for each drug class.

### Study population and exposure definitions

The study population comprised adult patients (aged ≥ 18 years at diagnosis) with a confirmed RCC diagnosis in the tumor registry who had at least one prescription for a VEGFR-TKIs recorded in the PRESCRIBING table between September 21, 2009, and June 22, 2023. The index date was defined as the date of the first VEGFR-TKIs prescription. Patients were excluded if they had no confirmed RCC diagnosis or a negative recorded OS time. Patients were classified into two mutually exclusive treatment arms: the experimental arm (EXP), comprising patients who received both a VEGFR-TKIs and a PD-1/PD-L1 inhibitor at any point during the study period; and the control arm (CTRL), comprising patients who received a VEGFR-TKIs with no documented PD-1/PD-L1 inhibitor exposure. VEGFR-TKIs include: sorafenib, sunitinib, pazopanib, axitinib, regorafenib, cabozantinib, lenvatinib, nintedanib, tivozanib, fruquintinib and vandetanib. PD-1/PD-L1 inhibitors include: nivolumab, pembrolizumab, atezolizumab, durvalumab, avelumab, cemiplimab, cosibelimab, dostarlimab, tislelizumab, toripalimab, and retifanlimab. Each agent was identified using RxNorm concept unique identifiers (RxCUI) through standardized reference tables. **(Supplementary Table 1)** Patients were assigned to the EXP arm if they had at least one prescription for an agent from each class; all remaining VEGFR-TKIs users with no recorded PD-1/PD-L1 inhibitor exposure were assigned to the CTRL arm.

### Covariates and Outcomes

Thirteen baseline covariates were identified as potential confounders of the association between treatment and outcome, including nine comorbidity variables and four demographic or behavioral variables. **(Supplementary Table 2)** Comorbidities were selected based on the NCI Comorbidity Index, an adaptation of the Charlson framework optimized for administrative data and widely used in oncology outcomes research^27^. The NCI index includes 13 comorbidity domains: myocardial infarction (MI), congestive heart failure (CHF), peripheral vascular disease (PVD), cerebrovascular disease (CVD), chronic obstructive pulmonary disease (COPD), dementia, paralysis (PARA), diabetes mellitus, chronic kidney disease (CKD), liver disease (LD), peptic ulcer disease (PUD), rheumatologic disease, and AIDS.^28^ Four conditions (dementia, PUD, rheumatologic disease, and AIDS) were excluded from the final analysis because fewer than 10 patients in the cohort carried these diagnoses, limiting their analytical stability. The final comorbidity set, therefore, included MI, CHF, PVD, CVD, COPD, PARA, diabetes mellitus, CKD, and LD. Comorbidities were identified from the complete diagnostic history recorded in the DIAGNOSIS table without restriction on the look-back period and were coded as binary indicators of presence or absence. Demographic and behavioral covariates included age at the index date (continuous), sex (binary), ethnicity (non-Hispanic Black, binary), and smoking history (binary). The primary outcome was overall survival (OS), defined as the time in months from the index date to the date of death recorded in the DEATH table from the Tumor Registry or to administrative censoring at 24 months, whichever occurred first. Patients alive at 24 months were censored, with OS time set to 24 months and event status coded as 0.

### Statistical analysis

Propensity score matching. To reduce confounding and approximate a randomized comparison, propensity scores were estimated using a logistic regression model with treatment assignment (COHORT_EXP: 1 = EXP, 0 = CTRL) as the dependent variable and all prespecified baseline covariates as predictors. Matching was performed using nearest-neighbor algorithms implemented in the MatchIt package in R (version 4.3.3). A variable ratio matching scheme was applied, allowing up to two control individuals to be matched to each treated individual (minimum 1), without replacement. A caliper width of 0.2 standard deviations of the logit of the propensity score was imposed, consistent with established recommendations^26^ to balance bias reduction and sample retention. This variable ratio approach was chosen to maximize the number of matched subjects while maintaining comparability between groups under a fixed caliper constraint. Individuals without eligible matches within the specified caliper were excluded from the matched sample. Covariate balance was evaluated using standardized mean differences for each variable, with values below 0.10 indicating adequate balance^29^. Balance diagnostics were visualized using Love plots generated with the cobalt package.

Kaplan-Meier analysis. Overall survival in the propensity score-matched cohort was compared by treatment group using the Kaplan-Meier method. Matching weights were incorporated into the survival estimates to account for the variable-ratio matching scheme. Median survival time and survival probabilities at 12 and 24 months were reported with corresponding 95% confidence intervals. Differences between groups were assessed using the weighted Cox model described below. The Kaplan-Meier estimator provides a nonparametric estimate of the survival function in the presence of censored data, while the *P*-value derived from the robust Cox model reflects the treatment effect after accounting for the matched study design.

Cox proportional hazards model. The primary effect estimate was the hazard ratio derived from a Cox proportional hazards model with treatment group as the sole covariate. To account for within-subclass correlation induced by the propensity score matching procedure, standard errors were estimated using a cluster-robust variance estimator, with clustering defined at the level of the matched subclass. This approach yields valid inference under correlated observations within matched sets. Hazard ratios are reported with 95% confidence intervals and corresponding two-sided *P*-values.

Proportional hazards assumption and RMST sensitivity analysis. The proportional hazards (PH) assumption was assessed using scaled Schoenfeld residuals via the cox.zph function in the survival package. Because treatment effects with VEGFR-TKIs plus PD-1/PD-L1 inhibitors combination therapy may not satisfy the proportional hazards assumption, RMST was pre-specified to provide an additional measure of treatment effect alongside the primary weighted Kaplan– Meier and Cox analyses. Standard RMST implementations do not accommodate the variable-ratio matching weights, so RMST was estimated directly from the weighted Kaplan–Meier survival curves as the area under the curve up to a truncation time of τ = 24 months, incorporating the same matching weights used in the primary analyses. Variance was computed using a weighted Greenwood estimator, and between-group differences and ratios, together with the corresponding 95% confidence intervals and two-sided *P*-values, were obtained via Wald tests on the additive and log scales, respectively. RMST provides an alternative, model-free summary of survival experience by quantifying the area under the survival curve up to a fixed time horizon. It can be interpreted as the average survival time within the first τ months and does not rely on the PH assumption, making it particularly suitable in settings with delayed treatment effects or crossing hazards.

All data preprocessing was performed in Python3 using the pandas, numpy, and lifelines libraries. All statistical analyses were conducted in R (version 4.3.3) using MatchIt, survival, survminer, cobalt, tableone, and ggplot2. All tests were two-sided, with statistical significance defined as *P* < 0.05.

## DATA AVAILABILITY

The data used in this study were obtained from the UF Health Integrated Data Repository (IDR). Due to institutional and patient privacy restrictions, the data is not publicly available. Access to the data may be granted upon reasonable request and with appropriate institutional approvals. Further information can be obtained from the corresponding author.

## AUTHOR CONTRIBUTIONS

D.S. and Q.S. were responsible for the implementation of all components of the study, including data cleaning and preprocessing, cohort construction, the overall survival analysis, and the drafting of the main body of the manuscript. J.S. conceived and designed the study, provided oversight of the overall analytical framework, and contributed to refinement of the study design and to manuscript drafting and revision. X.L. provided methodological and statistical guidance for the overall survival analysis, including supervision and validation of the statistical approaches, and contributed to manuscript drafting and revision. Y.C. (Yao Chen) provided statistical input, guided the literature search for relevant trials, and performed the manuscript revisions. All authors reviewed and approved the final version of the manuscript.

## CONFLICT OF INTEREST DISCLOSURES

The authors have no conflict of interest to disclose.

## FUNDING SUPPORT

Q.S. is supported by the National Institute of General Medical Sciences of the National Institutes of Health (R35GM151089). J.S. is supported by the National Library of Medicine of the National Institutes of Health (R01LM013771). J.S. is also supported by the National Institute of Health Office of the Director (OT2OD031919), the Indiana University Melvin and Bren Simon Comprehensive Cancer Center Support Grant from the National Cancer Institute (P30CA 082709), and the Indiana University Precision Health Initiative.

## Notes

### Competing Interest Statement

The authors have declared no competing interest.

### Author Declarations

Ethical approval was granted by the University of Florida Institutional Review Board (IRB202601021).

### Summary of Updates

The manuscript has been updated to reflect the current IRB approval number. No other substantive changes have been made.

