## Supplemental Information for "Combining VEGFR tyrosine kinase inhibitors and PD-1/PD-L1 inhibitors versus VEGFR tyrosine kinase inhibitors monotherapy in renal cell carcinoma: a target trial emulation" (from another version of this preprint: version 1, 2026-07-02)

**SUPPLEMENTARY INFORMATION**

**Supplementary Table 1. RxNorm concept codes (RxCUI) used to identify drug exposure for the study cohort. RxNorm** concept unique identifiers (RxCUIs) used to define exposure to study drugs in the UF Health Integrated Data Repository, including PD-1 and PD-L1 immune checkpoint inhibitors (ICIs) and VEGFR tyrosine kinase inhibitors (TKIs). Drugs are grouped by mechanism class, and all RxCUIs corresponding to each agent were included to ensure complete capture of prescribing records.

| **Drug Name** | **RxNorm RxCUI** |
| --- | --- |
| PD-L1 inhibitors |  |
| Atezolizumab | 1792781, 2693894, 1792776, 2693887, 2693883, 1792785, 2119696, 2693899, 1792782, 2693895, 1792783, 2693897, 2693898, 1792784, 2693896, 1792780, 2119695, 2693892, 1792777, 2693884, 1792779, 2693889, 1792778, 2693888 |
| Avelumab | 1875543, 1875534, 1875547, 1875544, 1875545, 1875546, 1875542, 1875539, 1875541, 1875540 |
| Cosibelimab | 2717520, 2717511, 2717512, 2717525, 2717521, 2717523, 2717524, 2717522, 2717518, 2717513, 2717515, 2717514 |
| Durvalumab | 1919508, 1919503, 1919512, 1919516, 1919509, 1919510, 1919511, 1919507, 1919515, 1919504, 1919506, 1919505 |
| PD-1 inhibitors |  |
| Cemiplimab | 2058831, 2058826, 2058825, 2058835, 2058832, 2058833, 2657678, 2058834, 2058830, 2058827, 2058829, 2058828 |
| Dostarlimab | 2539973, 2539967, 2539968, 2539977, 2539974, 2539975, 2656339, 2539976, 2539972, 2539969, 2539971, 2539970 |
| Nivolumab | 1597881, 2596780, 2700800, 1597876, 2596775, 2700793, 2596770, 2700791, 1657192, 1657196, 1991413, 2569081, 2596784, 2700805, 1597882, 2596781, 2700801, 1657191, 2596783, 2700803, 2654690, 2700804, 1597884, 2596782, 2700802, 1657190, 1657195, 1991412, 2569080, 2596778, 2700798, 1597877, 2596771, 2700792, 1657189, 2596777, 2700795, 1597878, 2596776, 2700794 |
| Pembrolizumab | 1547550, 2723764, 1547545, 2723757, 2723753, 1657751, 2723769, 2723772, 1547551, 2723765, 1657748, 2723767, 2723768, 1547553, 2723766, 1657746, 1657750, 2723762, 2723771, 1547546, 1657744, 2723754, 1657745, 2723759, 1547547, 2723758 |
| Retifanlimab | 2632987, 2632981, 2632980, 2632991, 2632988, 2632990, 2654692, 2632989, 2632985, 2632982, 2632984, 2632983 |
| Tislelizumab | 2677434, 2677426, 2677425, 2677439, 2677435, 2677437, 2677438, 2677436, 2677432, 2677427, 2677429, 2677428 |
| Toripalimab | 2669415, 2669406, 2669407, 2669420, 2669416, 2669418, 2669419, 2669417, 2669413, 2669408, 2669410, 2669409 |
| VEGFR tyrosine kinase inhibitors |  |
| Axitinib | 1243005, 1242999, 1243010, 1243014, 1243006, 1243013, 1243007, 1243008, 1243009, 1243004, 1243012, 1243000, 1243011, 1243003, 1243001, 1243002 |
| Cabozantinib | 1363274, 1790162, 1363408, 1363410, 1364581, 1363312, 1363409, 1364580, 1363268, 1363267, 1363279, 1363284, 1790167, 1790171, 1790175, 1363275, 1363283, 1790163, 1790170, 1790174, 1363276, 1790164, 1363277, 1363278, 1790165, 1790166, 1363273, 1363281, 1790161, 1790169, 1790173, 1363269, 1363280, 1790168, 1790172, 1363272, 1790160, 1363270, 1363271 |
| Fruquintinib | 2670185, 2670179, 2670190, 2670198, 2670186, 2670197, 2670189, 2670187, 2670188, 2670184, 2670196, 2670180, 2670195, 2670183, 2670181, 2670182 |
| Lenvatinib | 1792372, 1604350, 1604352, 1604384, 1604395, 1792393, 1796665, 2054157, 2054159, 1604349, 1604351, 1604383, 1604394, 1792392, 1796664, 2054156, 2054158, 1603296, 1604346, 1792377, 1796538, 1792373, 1796537, 1792374, 1792375, 1792376, 1604345, 1604348, 1604341, 1604347, 1604344, 1604342, 1604343 |
| Nintedanib | 1592743, 1592737, 1592736, 1592748, 1592752, 1592744, 1592751, 1592745, 1592746, 1592747, 1592742, 1592750, 1592738, 1592749, 1592741, 1592739, 1592740 |
| Pazopanib | 867503, 714438, 732187, 867506, 867504, 867505, 1187595, 1187596, 867502, 867508, 867500, 867507, 867501, 1161365, 1161366 |
| Regorafenib | 1312403, 1312397, 1371319, 1312408, 1312404, 1312405, 1312406, 1312407, 1312402, 1312398, 1312401, 1312399, 1312400 |
| Sorafenib | 619985, 495881, 597744, 615979, 615977, 615978, 1182862, 1182863, 597747, 597745, 597746, 1159691, 1159692 |
| Sunitinib | 616274, 357977, 616275, 1541928, 616283, 616287, 616292, 1541927, 616281, 616286, 616291, 616282, 1185474, 1185475, 1541890, 616279, 616285, 616289, 1541889, 616277, 616284, 616288, 616278, 1160852, 1160853 |
| Tivozanib | 2534239, 2534233, 2534245, 2534244, 2534249, 2534240, 2534248, 2534241, 2534242, 2534243, 2534238, 2534247, 2534234, 2534246, 2534237, 2534235, 2534236 |
| Vandetanib | 1114071, 1098413, 1114074, 1114096, 1114072, 1114095, 1114073, 1172982, 1173663, 1098416, 1098418, 1098414, 1098417, 1098415, 1160503, 1160504 |

**Supplementary Table 2. ICD-9 and ICD-10 diagnosis codes used to define baseline comorbidities.** ICD-9 and ICD-10 diagnosis codes used to identify baseline comorbidities in the UF Health Integrated Data Repository. For each condition, codes from the National Cancer Institute (NCI) 2021 comorbidity definition are shown alongside the operational definition applied in this study ("Our Definition"); the Notes column documents how the two differ. Where the NCI separates acute and historical events, our definition combines them into a single condition. For several conditions, our definition uses code stems or a subset of the NCI codes rather than the full NCI list, as noted. "x" denotes a wildcard matching all sub-codes within the stated category.

| **Condition** | **NCI 2021 Definition** | | **Condition** | **Our Definition** | | **Notes** |
| --- | --- | --- | --- | --- | --- | --- |
|  | **ICD-9** | **ICD-10** |  | **ICD-9** | **ICD-10** |  |
| **Acute Myocardial Infarction** | 410.x | I21.x, I22.x | **Myocardial Infarction** | 410.x, 412.x | I21.x, I22.x, I25.2 | *MI includes History of MI codes (412, I25.2) and Acute MI codes (410, I21, I22)* |
| **History of Myocardial Infarction** | 412.x | I25.2 |  |  |  |  |
| **Congestive Heart Failure** | 398.91, 402.01, 402.11, 402.91, 404.01, 404.03, 404.11, 404.13, 404.91, 404.93, 425.4–425.9, 428.x | I09.9, I11.0, I13.0, I13.2, I25.5, I42.0, I42.5–I42.9, I43.x, I50.x, P29.0 | **Congestive Heart Failure** | 428.x | I50.x | *Our definition uses stems only; NCI has a broader list* |
| **Peripheral Vascular Disease** | 093.0, 440.x, 441.x, 443.1–443.9, 447.1, 557.1, 557.9, V43.4 | I70.x, I71.x, I73.1, I73.8, I73.9, I77.1, I79.0, I79.2, K55.1, K55.8, K55.9, Z95.8, Z95.9 | **Peripheral Vascular Disease** | 443.x, 440.x | I73.x | *Our definition is a subset of NCI (I73, 440, 443)* |
| **Cerebrovascular Disease** | 362.34, 430.x–438.x | G45.x, G46.x, H34.0, I60.x–I69.x | **Cerebrovascular Disease** | 362.34, 430.x–438.x | G45.x, G46.x, H34.0, I60.x–I69.x |  |
| **Chronic Obstructive Pulmonary Disease** | 416.8, 416.9, 490.x–505.x, 506.4, 508.1, 508.8 | I27.8, I27.9, J40.x–J47.x, J60.x–J67.x, J68.4, J70.1, J70.3 | **Chronic Obstructive Pulmonary Disease** | 491.x, 492.x, 496.x | J44.x | *Our definition is a subset (J44, 491–496)* |
| **Dementia** | 290.x, 294.1, 331.2 | F00.x–F03.x, F05.1, G30.x, G31.1 | **Dementia** | 290.x, 294.1, 331.2 | F00.x–F03.x, F05.1, G30.x, G31.1 |  |
| **Paralysis** | 334.1, 342.x, 343.x, 344.0–344.6, 344.9 | G04.1, G11.4, G80.1, G80.2, G81.x, G82.x, G83.0–G83.4, G83.9 | **Paralysis** | 334.1, 342.x, 343.x, 344.0–344.6, 344.9 | G04.1, G11.4, G80.1, G80.2, G81.x, G82.x, G83.0–G83.4, G83.9 |  |
| **Diabetes (uncomplicated)** | 250.0–250.3, 250.8, 250.9 | E10.0, E10.1, E10.6, E10.8, E10.9, E11.0, E11.1, E11.6, E11.8, E11.9, E13.0, E13.1, E13.6, E13.8, E13.9 | **Diabetes** | 250.x | E10.x, E11.x, E13.x | *Our definition uses stems (250, E10/11/13); covers both uncomplicated and with complications* |
| **Diabetes with Complications** | 250.4–250.7 | E10.2–E10.5, E10.7, E11.2–E11.5, E11.7, E13.2–E13.5, E13.7 |  |  |  |  |
| **Renal Disease** | 403.01, 403.11, 403.91, 404.02, 404.03, 404.12, 404.13, 404.92, 404.93, 582.x, 583.0–583.7, 585.x, 586.x, 588.0, V42.0, V45.1, V56.x | I12.0, I13.1, N03.2–N03.7, N05.2–N05.7, N18.x, N19.x, N25.0, Z49.0–Z49.2, Z94.0, Z99.2 | **Kidney Disease** | 403.01, 403.11, 403.91, 404.02, 404.03, 404.12, 404.13, 404.92, 404.93, 582.x, 583.0–583.7, 585.x, 586.x, 588.0, V42.0, V45.1, V56.x | I12.0, I13.1, N03.2–N03.7, N05.2–N05.7, N18.x, N19.x, N25.0, Z49.0–Z49.2, Z94.0, Z99.2 |  |
| **Mild Liver Disease** | 070.22, 070.23, 070.32, 070.33, 070.44, 070.54, 070.6, 070.9, 570.x, 571.x, 573.3, 573.4, 573.8, 573.9, V42.7 | B18.x, K70.0–K70.3, K70.9, K71.3–K71.5, K71.7, K73.x, K74.x, K76.0, K76.2–K76.4, K76.8, K76.9, Z94.4 | **Liver Disease** | 070.22, 070.23, 070.32, 070.33, 070.44, 070.54, 070.6, 070.9, 570.x, 571.x, 573.3, 573.4, 573.8, 573.9, V42.7,  456.0–456.2, 572.2–572.8 | B18.x, K70.0–K70.3, K70.9, K71.3–K71.5, K71.7, K73.x, K74.x, K76.0, K76.2–K76.4, K76.8, K76.9, Z94.4,  I85.0, I85.9, I86.4, I98.2, K70.4, K71.1, K72.1, K72.9, K76.5, K76.6, K76.7 | *LD variable combines Mild + Moderate/Severe Liver Disease* |
| **Moderate/Severe Liver Disease** | 456.0–456.2, 572.2–572.8 | I85.0, I85.9, I86.4, I98.2, K70.4, K71.1, K72.1, K72.9, K76.5, K76.6, K76.7 |  |  |  |  |
| **Peptic Ulcer Disease** | 531.x–534.x | K25.x–K28.x | **Peptic Ulcer Disease** | 531.x–534.x | K25.x–K28.x |  |
| **Rheumatologic Disease** | 446.5, 710.0–710.4, 714.0–714.2, 714.8, 725.x | M05.x, M06.x, M31.5, M32.x–M34.x, M35.1, M35.3, M36.0 | **Rheumatologic Disease** | 446.5, 710.0–710.4, 714.0–714.2, 714.8, 725.x | M05.x, M06.x, M31.5, M32.x–M34.x, M35.1, M35.3, M36.0 |  |
| **AIDS** | 042.x–044.x | B20.x–B22.x, B24.x | **AIDS** | 042.x–044.x | B20.x–B22.x, B24.x |  |
